# Associations of Telemedicine vs. In-Person Ambulatory Care Visits on Cancellation Rates and 30-Day Follow-Up Hospitalizations and Emergency Department Visits

**DOI:** 10.1101/2021.08.13.21262018

**Authors:** Julianne N. Kubes, Ilana Graetz, Zanthia Wiley, Nicole Franks, Ambar Kulshreshtha

**Affiliations:** Office of Quality and Risk, Emory Healthcare; Department of Health Policy and Management, Rollins School of Public Health, Emory University; Division of Infectious Diseases, Emory University School of Medicine; Department of Emergency Medicine, Emory University School of Medicine; Division of Family and Preventative Medicine, Emory University School of Medicine; Department of Epidemiology, Rollins School of Public Health, Emory University

## Abstract

**Importance:** Studies have shown that telemedicine use in specific conditions can promote continuity of care, decreases healthcare costs, and can potentially improve clinical outcomes. The COVID-19 pandemic forced many healthcare systems to expand access for patients using telemedicine, but little is known about cancellation frequencies in telemedicine vs. in-person appointments and its impact on clinical outcomes.

**Objective:** Compare ambulatory clinic cancellation rates, 30-day inpatient hospitalizations rates, and 30-day emergency department visit rates between in-person and video telemedicine appointments, and examine differences in cancellation rates by age, race/ethnicity, gender, and insurance.

**Design:** A retrospective cohort study.

**Setting:** The largest academic healthcare system in the state of Georgia with ambulatory clinics in urban, suburban and rural settings.

**Participants:** Adults scheduled for an ambulatory clinic appointment from June 2020 to December 2020 were included. Each appointment was identified as either a video telemedicine or in-person clinic appointment. Demographics including age, race, ethnicity, gender, primary insurance, and comorbidities were extracted from the electronic medical record.

**Main Outcomes and Measures:** The primary process outcome was ambulatory clinic cancellation rates. The primary clinical outcomes were 30-day hospitalization rates and 30-day emergency department visit rates. Multivariable logistic regression was used to assess differences in the clinical outcomes between appointment types.

**Results:** A total of 1,652,623 ambulatory clinic appointments were scheduled during the study period. Ambulatory appointment cancellations rates were significantly lower among telemedicine appointments compared to in-person appointments (20.5% vs. 31.0%, p <.001). Cancellation rates were significantly lower for telemedicine appointments than in-person appointments regardless of gender, age, race, ethnicity, primary insurance, or specialty (p <.05 for all sub-groups). Telemedicine appointments was associated with lower 30-day hospitalization rates compared to in-person appointments (2.1% vs. 2.8%; aOR: 0.72, 95% CI: 0.71 to 0.74). There was no difference in 30-day emergency department visit rates between telemedicine and in-person appointment patients (2.6% vs. 2.6%: aOR: 1.00, 95% CI: 0.98 to 1.02)

**Conclusions and Relevance:** Our findings suggest that there are fewer barriers to attending an ambulatory care visit via telemedicine relative than in-person. Moreover, using telemedicine was not associated with any more frequent adverse clinical events compared with in-person visits.

## Introduction

In response to the COVID-19 pandemic, several healthcare systems rapidly developed telemedicine programs to provide ongoing access for patients to receive ambulatory care from their regular providers.^1^ Rapid legislative and regulatory changes to payment and privacy requirements supported the expansion of telemedicine in United States.^2^ Prior studies have shown that telemedicine can safely and effectively be used to diagnose and treat acute conditions, monitor chronic conditions, conduct follow-up visits while reducing travel and wait times for patients, and potentially lower healthcare costs.^3-6^ There is, however, limited data on telemedicine visits compared with in-person visits in regards to healthcare access and outcomes such as preventing emergency department (ED) visits and subsequent hospitalizations.

Telemedicine expansion presents an opportunity to generate further evidence regarding its effectiveness and utility and whether it helps to improve patient outcomes.^7^ Our study is one of the first large scale studies, encompassing a diverse physician and patient population, to look at ambulatory clinic cancellations, hospitalizations, and ED outcomes in a major academic healthcare system. We hypothesize that compared with in-person visits, telemedicine visits have lower cancellation rates regardless of age, gender, and race of the patient, and similar rates of hospitalizations and ED visits.

## Methods

Emory Healthcare (EHC) is the largest healthcare system in the state of Georgia with more than 2800 physicians and 250 provider locations in urban, suburban and rural settings. EHC began using Zoom (Zoom Video Communication, Inc., San Jose, CA) in April 2020 to offer telemedicine appointments for ambulatory care visits and included both an audio and video component in a synchronous format. Our study included adult patients (age ≥ 18 years) scheduled for an ambulatory clinic appointment between June and December 2020 within EHC (when patients had a choice of in-person or telemedicine visit). We selected the above study period because appointments made in March-May 2020 were intentionally assigned to telemedicine due to the COVID-19 surge in Georgia. We defined a telemedicine appointment as an appointment conducted via video and an in-person appointment as an appointment conducted at an ambulatory clinic when the patient was physically present. Demographic information was extracted from the electronic medical record including age, race, ethnicity, gender, primary insurance, and comorbidities identified by billed ICD-10 diagnosis codes. A Charlson Comorbidity Index (CCI) was calculated for each patient for risk adjustment.^8^ We excluded patients with a positive SARS-Co-V-2 (COVID-19) PCR test within 14 days of their appointment, as these appointments were intentionally assigned to telemedicine. No patient identifiers were used, and the study was reviewed and deemed exempt by Emory Institutional Review Board review.

The primary process outcome was ambulatory clinic cancellation rates, defined as the percentage of ambulatory clinic appointments where the patient cancelled beforehand or did not show to the appointment. The primary clinical outcomes were 30-day hospitalization and ED rates, defined as the percentage of ambulatory patients who were admitted as an inpatient to a hospital or had an ED visit within 30 days of their ambulatory appointment.

Differences in cancellation rates between telemedicine and in-person appointments and among sub-groups were compared using the Chi-square test. Multivariable logistic regression was used to compare 30-day hospitalization and ED visit rates between telemedicine and in-person appointments, adjusting for age and CCI. Statistical analyses were performed using R (version 4.0.2; Rstudio, Inc., Boston, MA). This study followed the Strengthening the Reporting of Observational Studies in Epidemiology (<u>STROBE</u>) reporting guideline.^9^

## Results

A total of 1,652,623 ambulatory clinic appointments were scheduled during the study timeframe and met the inclusion criteria. Of those, 412,936 (25.0%) were telemedicine appointments and 1,239,687 (75.0%) were in-person appointments. The average age was 59 years (SD 18.4), 61.1% were female, 47.5% were White, and 35.8% were Black (Table 1).

**Table 1.** Telemedicine and In-Person Outpatient Appointment Patient Characteristics, June – December 2020

| **Patient Characteristic** | **Telemedicine**<br>**Appointments**<br>**(n = 412,936)** | **In-Person**<br>**Appointments**<br>**(n = 1,239,687)** |
| --- | --- | --- |
| Female, No. (%) | 258,761 (62.7) | 750,419 (60.5) |
| Age, No. (%) |  |  |
| < 18 | 11,722 (2.8) | 29,835 (2.4) |
| 18-34 | 56,946 (13.8) | 147,961 (11.9) |
| 35-64 | 207,552 (50.3) | 575,966 (46.5) |
| 65+ | 136,716 (33.1) | 485,928 (39.2) |
| Race, No. (%) |  |  |
| White | 199,653 (48.3) | 585,060 (47.2) |
| Black | 147,058 (35.6) | 444,218 (35.8) |
| Asian | 11,604 (2.8) | 38,761 (3.1) |
| American Indian/Alaska Native | 940 (0.2) | 3,315 (0.3) |
| Native Hawaiian/Pacific Islander | 776 (0.2) | 2,911 (0.2) |
| Multiple | 2,425 (0.6) | 6,628 (0.5) |
| Unknown | 50,480 (12.2) | 185,794 (12.8) |
| Ethnic Group, No. (%) |  |  |
| Non-Hispanic | 218,074 (77.0) | 949,663 (76.6) |
| Hispanic | 10,862 (2.6) | 34,309 (2.8) |
| Unknown | 84,000 (20.3) | 255,718 (20.6) |
| Primary Insurance, No. (%) |  |  |
| Commercial | 216,626 (52.5) | 561,236 (45.3) |
| Medicare | 147,864 (35.8) | 486,933 (39.3) |
| Medicaid | 21,674 (5.2) | 58,233 (4.7) |
| Uninsured | 26,772 (6.5) | 133,285 (10.7) |
| Specialty, No. (%) |  |  |
| Primary Care | 131,353 (31.8) | 359,537 (29.0) |
| Sub-Specialty | 254,058 (61.5) | 785,511 (63.4) |
| Surgical | 27,525 (6.7) | 94,642 (7.6) |
| Comorbidities, No. (%) |  |  |
| Hypertension | 214,669 (52.0) | 671,601 (54.2) |
| Acute myocardial infarction | 18,020 (4.4) | 67,055 (5.4) |
| Congestive heart failure | 46,692 (11.3) | 171,228 (13.8) |
| Dementia | 13,385 (3.2) | 26,823 (2.2) |
| Diabetes | 94,199 (22.8) | 295,017 (23.8) |
| Chronic obstructive pulmonary disease | 82,217 (19.9) | 239,311 (19.3) |
| Renal disease | 59,640 (14.4) | 186,501 (15.0) |
| Malignancy | 70,149 (17.0) | 262,362 (21.2) |
| Charlson Comorbidity Index, No. (%) |  |  |
| 0-1 (low risk) | 266,945 (64.6) | 774,433 (62.5) |
| 2-3 (medium risk) | 93,389 (22.6) | 294,212 (23.7) |
| 4+ (high risk) | 52,602 (12.7) | 171,045 (13.8) |

Ambulatory appointment cancellation rates were significantly lower among telemedicine appointments compared to in-person appointments (20.5% vs. 31.0%, p <.0001, Table 2). Cancellation rates were lower for telemedicine regardless of gender, age, race, ethnicity, primary insurance, or specialty (p <.05 for all sub-groups, Table 2).

**Table 2.** Telemedicine and In-Person Outpatient Cancellations, June – December 2020.

| **Sub-Group** | **Telemedicine**<br>**Appointments**<br>**(n = 412,936)** | **In-Person**<br>**Appointments**<br>**(n = 1,239,687)** | **P value** |
| --- | --- | --- | --- |
| Total cancellations, No. (%) | 84,211 (20.4) | 383,902 (31.0) | <.0001 |
| Gender, No. (%) |  |  |  |
| Male | 31,232 (20.3) | 145,613 (29.8) | <.0001 |
| Female | 25,979 (20.5) | 238,267 (31.8) | <.0001 |
| Age, No. (%) |  |  |  |
| < 18 | 2,251 (19.2) | 9,015 (30.2) | <.0001 |
| 18-34 | 11,157 (19.6) | 44,546 (30.1) | <.0001 |
| 35-64 | 41,651 (20.1) | 100,012 (31.3) | <.0001 |
| 65+ | 29,152 (21.3) | 150,329 (30.9) | <.0001 |
| Race, No. (%) |  |  |  |
| White | 29,383 (19.7) | 181,464 (31.0) | <.0001 |
| Black | 30,133 (20.5) | 135,505 (30.5) | <.0001 |
| Asian | 2,571 (22.2) | 118,56 (30.6) | <.0001 |
| American Indian/Alaska Native | 199 (21.2) | 1,007 (30.4) | <.0001 |
| Native Hawaiian/Pacific Islander | 167 (21.5) | 853 (29.3) | <.0001 |
| Multiple | 493 (20.3) | 2,110 (31.8) | <.0001 |
| Ethnic Group, No. (%) |  |  |  |
| Non-Hispanic | 63,916 (20.1) | 292,156 (30.8) | <.0001 |
| Hispanic | 2,442 (22.5) | 10,204 (29.7) | <.0001 |
| Insurance, No. (%) |  |  |  |
| Commercial | 38,521 (17.8) | 152,115 (27.1) | <.0001 |
| Medicare | 28,978 (19.6) | 140,695 (28.9) | <.0001 |
| Medicaid | 4,002 (18.5) | 16,656 (28.6) | <.0001 |
| Uninsured | 3,414 (24.5) | 17,419 (25.3) | .04 |
| Specialty, No. (%) |  |  |  |
| Primary Care | 23,773 (18.1) | 115,192 (32.0) | <.0001 |
| Sub-Specialty | 54,326 (21.4) | 437,748 (30.3) | <.0001 |
| Surgical | 6,112 (22.2) | 30,962 (32.7) | <.0001 |

Telemedicine visits were associated with lower 30-day hospitalization rate compared to in-person appointments (2.1% vs. 2.8%; OR: 0.73, 95% CI: 0.71 to 0.74); this result did not change after adjusting for comorbid conditions (aOR: 0.72, 95% CI: 0.71 to 0.74, Table 3). We did not find a statistically significant difference in 30-day ED visit rate between telemedicine and in-person appointments (2.6% vs. 2.6%: OR: 0.99, 95% CI: 0.96 to 1.01) after adjusting for comorbid conditions (aOR: 1.00, 95% CI: 0.98 to 1.02).

**Table 3.**
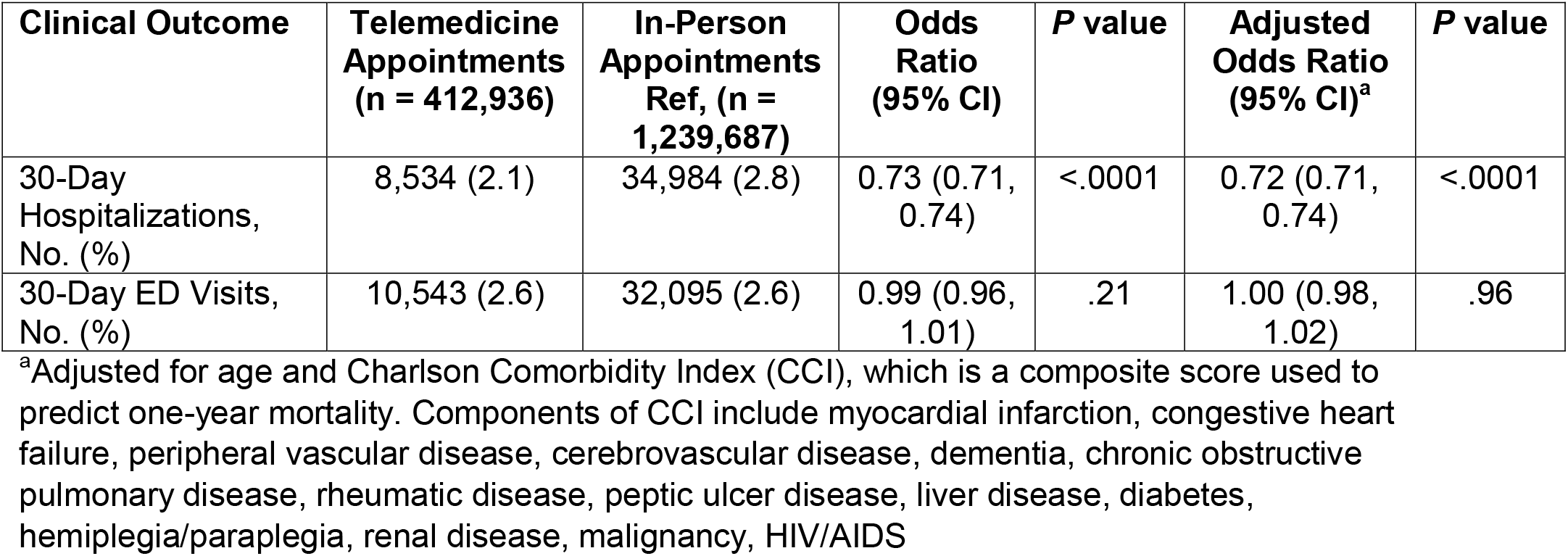
Crude and Risk-Adjusted Clinical Outcomes for Telemedicine vs. In-Person Outpatient Appointments, June – December 2020

| **Clinical Outcome** | **Telemedicine Appointments (n = 412,936)** | **In-Person Appointments Ref, (n = 1,239,687)** | **Odds Ratio (95% CI)** | **P value** | **Adjusted Odds Ratio (95% CI)<sup>a</sup>** | **P value** |
| --- | --- | --- | --- | --- | --- | --- |
| 30-Day Hospitalizations, No. (%) | 8,534 (2.1) | 34,984 (2.8) | 0.73 (0.71, 0.74) | <.0001 | 0.72 (0.71, 0.74) | <.0001 |
| 30-Day ED Visits, No. (%) | 10,543 (2.6) | 32,095 (2.6) | 0.99 (0.96, 1.01) | .21 | 1.00 (0.98, 1.02) | .96 |

<sup>a</sup>Adjusted for age and Charlson Comorbidity Index (CCI), which is a composite score used to predict one-year mortality. Components of CCI include myocardial infarction, congestive heart failure, peripheral vascular disease, cerebrovascular disease, dementia, chronic obstructive pulmonary disease, rheumatic disease, peptic ulcer disease, liver disease, diabetes, hemiplegia/paraplegia, renal disease, malignancy, HIV/AIDS

## Discussion

In a retrospective cohort study of adult patients receiving ambulatory care at a large academic healthcare system, telemedicine visits were associated with fewer cancellations than in-person visits during the COVID-19 pandemic and this was true for all population sub-groups. Moreover, using telemedicine was not associated with worse adverse clinical events, such as a follow-up ED visits or hospitalization.

Video telemedicine visits offer patients real-time and easier access directly with a clinician without leaving their homes. Prior concerns with telemedicine including barriers to technology use among older and minority populations also did not seem to hold true from our study.^10,11^ Our findings suggest that telemedicine may provide more convenient access to care and fewer barriers than in-person visits for most population sub-groups. Previous studies have found that travel and wait times in clinics continue to be a significant barrier to in-person care. ^12^Our findings suggest that these barriers may contribute to higher cancellation rates for in-person appointments. These barriers have also been found to be more common among minority groups, resulting in disparities in healthcare access.^13,14^ Convenience of telemedicine improves access to care, particularly among vulnerable patients.^15^ Our results add to the literature by showing that telemedicine appointments have a higher completion rate and is not associated with higher adverse clinical events within 30 days.

There are several limitations to the interpretation and generalizability of our findings. Our study was conducted at a single large healthcare system that used a common platform for most telemedicine visits during the COVID-19 pandemic. Our study is observational, and the results should not be interpreted as causal. Despite robust adjustment of patient risk factors, there is likely to be unmeasured confounding and our inability to know the reasons for patient cancellations. Finally, we relied on administrative and billing codes to capture visit information and there is potential for misclassification. However, it is likely to be non-differential and only bias the study results towards the null.

In conclusion, in a large academic health system, telemedicine appointments were cancelled significantly less than in-person appointments, regardless of age, race/ethnicity, gender, or insurance. Telemedicine appointments were associated with fewer 30-day hospitalizations compared to in-person appointments and had similar rates of ED visits. Telemedicine appointments increases access to healthcare patients from all social sub-groups and may help reduce healthcare disparities. Expansion of telemedicine in the US and globally warrants more efforts that focus on outcomes comparison to inform policy and clinical practice decisions. Future studies should examine differences in quality of care, patient clinical outcomes, and costs comparing telemedicine to in-person ambulatory visits more generally and for specific chronic conditions.

## Data Availability

All data was extracted from the electronic medical record via a secured and encrypted clinical data warehouse used at Emory Healthcare. Data is not available to individuals outside of Emory Healthcare or Emory University.

## Acknowledgements

We would like to acknowledge Gregory J. Esper, MD, MBA, Associate Chief Medical Officer at Emory Healthcare and Joel Shu, MD, Chief Medical Officer and Chief Quality Officer at Emory Healthcare Network for their insight and consult on this study. We would also like to acknowledge the CROSS Collaborative for their assistance with funding this study. All study authors declare no conflicts of interest.

## References

1. Patel SY, Mehrotra A, Huskamp HA, Uscher-Pines L, Ganguli I, Barnett ML. Trends in Outpatient Care Delivery and Telemedicine During the COVID-19 Pandemic in the US. JAMA Intern Med. 2021;181(3):388–391.

2. Mehrotra A, Bhatia RS, Snoswell CL. Paying for Telemedicine After the Pandemic. JAMA. 2021;325(5):431–432.

3. Zhang W, Cheng B, Zhu W, Huang X, Shen C. Effect of Telemedicine on Quality of Care in Patients with Coexisting Hypertension and Diabetes: A Systematic Review and Meta-Analysis. Telemed J E Health. 2020.

4. Dorsey ER, Topol EJ. State of Telehealth. N Engl J Med. 2016;375(14):1400.

5. Lin MH, Yuan WL, Huang TC, Zhang HF, Mai JT, Wang JF. Clinical effectiveness of telemedicine for chronic heart failure: a systematic review and meta-analysis. J Investig Med. 2017;65(5):899–911.

6. Zhai YK, Zhu WJ, Cai YL, Sun DX, Zhao J. Clinical-and cost-effectiveness of telemedicine in type 2 diabetes mellitus: a systematic review and meta-analysis. Medicine (Baltimore). 2014;93(28):e312.

7. Roberts ET, Mehrotra A. Assessment of Disparities in Digital Access Among Medicare Beneficiaries and Implications for Telemedicine. JAMA Intern Med. 2020;180(10):1386–1389.

8. Charlson ME, Pompei P, Ales KL, MacKenzie CR. A new method of classifying prognostic comorbidity in longitudinal studies: development and validation. J Chronic Dis. 1987;40(5):373–383.

9. Ghaferi AA, Schwartz TA, Pawlik TM. STROBE Reporting Guidelines for Observational Studies. JAMA Surg. 2021.

10. Eberly LA, Kallan MJ, Julien HM, et al. Patient Characteristics Associated With Telemedicine Access for Primary and Specialty Ambulatory Care During the COVID-19 Pandemic. JAMA Netw Open. 2020;3(12):e2031640.

11. Lam K, Lu AD, Shi Y, Covinsky KE. Assessing Telemedicine Unreadiness Among Older Adults in the United States During the COVID-19 Pandemic. JAMA Intern Med. 2020;180(10):1389–1391.

12. Ray KN, Chari AV, Engberg J, Bertolet M, Mehrotra A. Disparities in Time Spent Seeking Medical Care in the United States. JAMA Intern Med. 2015;175(12):1983–1986.

13. Shi L, Chen C-C, Nie X, Zhu J, Hu R. Racial and socioeconomic disparities in access to primary care among people with chronic conditions. The Journal of the American Board of Family Medicine. 2014;27(2):189–198.

14. Brown EJ, Polsky D, Barbu CM, Seymour JW, Grande D. Racial disparities in geographic access to primary care in Philadelphia. Health affairs. 2016;35(8):1374–1381.

15. Reed ME, Huang J, Graetz I, et al. Patient Characteristics Associated With Choosing a Telemedicine Visit vs Office Visit With the Same Primary Care Clinicians. JAMA network open. 2020;3(6):e205873–e205873.

